# Population-split-based risk assessment model of venous thromboembolism in Chinese medical inpatients

**DOI:** 10.1101/2022.01.08.22268955

**Authors:** Xin Wang, Yu-Qing Yang, Xin-Yu Hong, Si-Hua Liu, Jian-Chu Li, Ting Chen, Ju-Hong Shi

## Abstract

**Objective:** Inpatients with high risk of venous thromboembolism (VTE) usually face serious threats to their health and economic conditions. Many studies using machine learning (ML) models to predict VTE risk neglected an important statistical phenomenon, ‘fuzzy feature’, and achieved inferior results. Considering the effect of ‘fuzzy feature’, our study aims to develop a VTE risk assessment model suitable for Chinese medical inpatients.

**Materials and Methods:** Inpatients in the medical department of Peking Union Medical College Hospital (PUMCH) from January 2014 to June 2016 were collected. A new ML VTE risk assessment model was built through population splitting. First patients were classified into different groups based on values of VTE risk factors, then trustless groups were filtered out, and finally ML models were built on training data in unit of groups. Predictive performances of our method, five traditional ML models, and the Padua model were compared.

**Results:** The ‘fuzzy feature’ was verified on the whole dataset. Compared with the Padua model, the proposed model showed higher sensitivities and specificities on training data, and higher specificities and similar sensitivities on test data. Standard deviations of predictive validity of five ML models were larger than the proposed model.

**Discussion:** The proposed model was the only one which showed advantages on both sensitivity and specificity over Padua model. Its robustness was better than traditional ML models.

**Conclusion:** This study built a population-split-based ML model of VTE for Chinese medical inpatients and it may help clinicians stratify VTE risk and guide prevention more efficiently.

## INTRODUCTION

Venous thromboembolism (VTE), comprising deep venous thrombosis (DVT) and pulmonary embolism (PE), is a life-threatening disease associated with more than one-half million hospitalizations in the United States each year, and is a contributing cause in 100,000 or more deaths.^1,2^ As a common cardiovascular disease, VTE often leads to complications including recurrent VTE, post-pulmonary embolism syndrome, chronic thromboembolic pulmonary hypertension, and post-thrombotic syndrome, causing heavy burden to both life quality and economy. ^3^

Prophylaxis against VTE such as anticoagulant drugs, graduated compression stockings and venous foot pump can reduce mortality efficiently. Studies have shown that appropriate prevention can lower patients’ VTE incidence from 10.5%-14.9% to 5.5-5.6% in medical department and also reduce VTE events in surgical departments. ^4-6^ Since hypercoagulability is one of the most important VTE risk factors,^7^ anticoagulant drugs realize the disease prevention by changing coagulation status for high VTE risk patients. However, it may also cause bleeding events and even death, especially for patients with low VTE risk, who is not hypercoagulabe. Therefore, to recognize patients with high VTE risk clinically is vital and a precise VTE risk assessment model is needed to guide prevention.

The American College of Chest Physicians recommended Padua risk assessment model, a linear model consisting of 11 VTE risk factors (**Table 1)**, to stratify VTE risk for medical inpatients.^8,9^ Inpatients with Padua score of no less than 4 points were considered as high-risk and recommended to receive prophylaxis. However, due to the close correlations between VTE and race, genetic background and disease spectrum, studies have shown that the Padua model, which was derived based on the Western population, is not suitable for Chinese inpatients.^10,11^ Thus, it is necessary to establish a model suitable for Chinese inpatients.

**Table 1.**
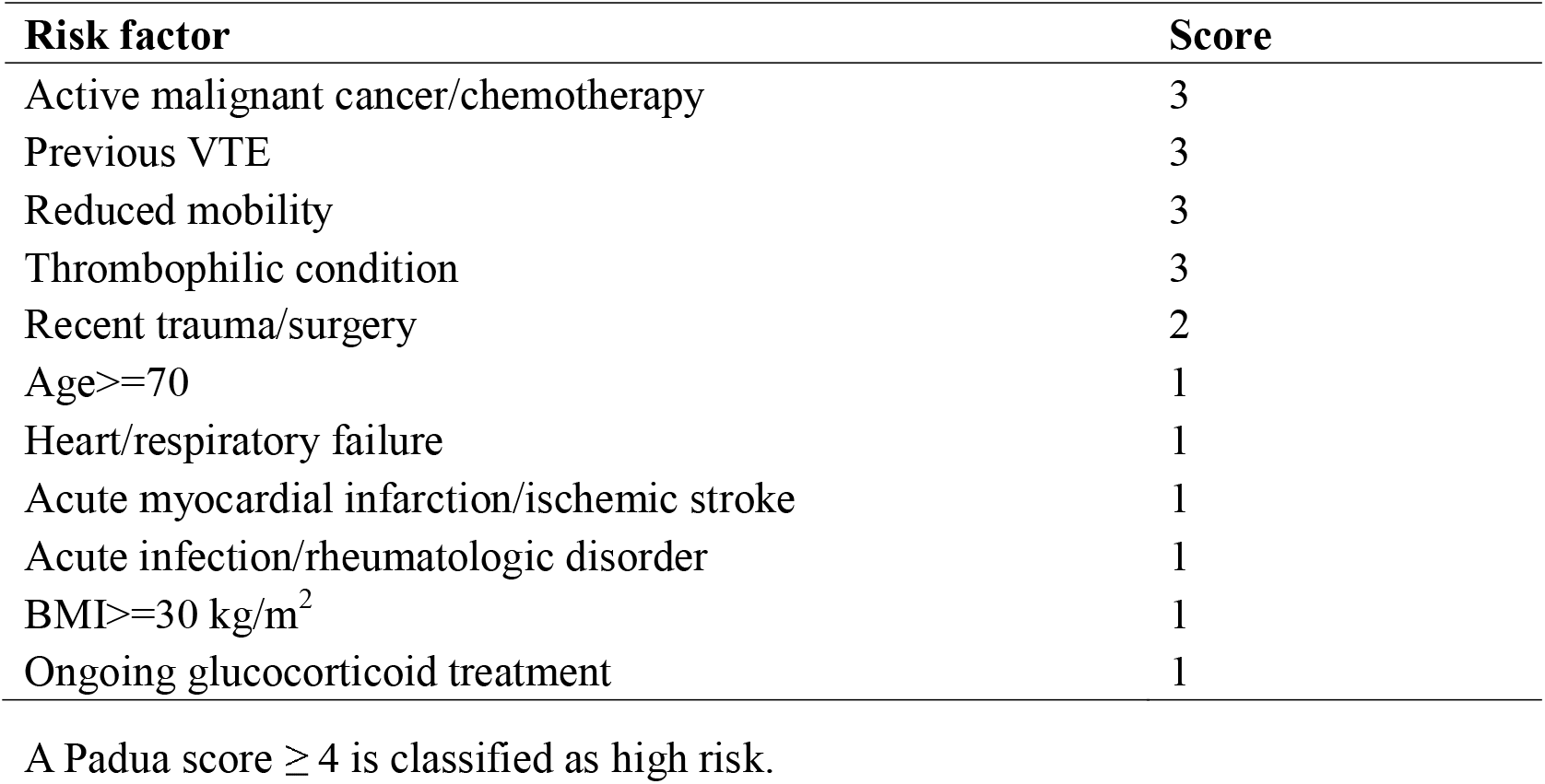
Padua risk assessment model.

| Risk factor | Score |
| --- | --- |
| Active malignant cancer/chemotherapy | 3 |
| Previous VTE | 3 |
| Reduced mobility | 3 |
| Thrombophilic condition | 3 |
| Recent trauma/surgery | 2 |
| Age$\geq 70$ | 1 |
| Heart/respiratory failure | 1 |
| Acute myocardial infarction/ischemic stroke | 1 |
| Acute infection/rheumatologic disorder | 1 |
| BMI$\geq 30$ kg/m<sup>2</sup> | 1 |
| Ongoing glucocorticoid treatment | 1 |

A Padua score $\geq 4$ is classified as high risk.

With the rapid development of artificial intelligence technology, machine learning (ML) models are increasingly employed in medical research.^12,13^ Support vector machine (SVM), random forest (RF), gradient boosting decision tree (GBDT), logistic regression (LR) and XGBoost have been proposed to do VTE risk assessment, but most of them trained models using equal number of VTE and non-VTE patients.^14-16^ Wang, Yang, Liu, Hong, Sun, Shi ^17^ compared multiple ML models by training them on 188 VTE and 188 non-VTE patients, and showed that performances of ML models were instable and their sensitivities were lower than the Padua. In addition, explainability of ensemble-based models such as RF, GBDT and XGBoost was limited though their predictive performances were relatively well.

All previous ML VTE risk assessment models ignored an important statistical phenomenon in VTE population (we called ‘fuzzy feature’). That is, VTE and non-VTE patients can share the same values of all risk factors. Let *x*_*i*_ = (*x*_*i*1_, *x*_*i*2_, *x*_*i*3_, …, *x*_*iK*_) be the *i*^*th*^ patient with K risk factors where *x*_*ij*_ ∈ {0,1} represents value of the *j*^*th*^ risk factor, and *y*_*i*_ ∈ {0,1} indicates whether the patient has VTE. The ‘fuzzy feature’ phenomenon means that there are the *i*^*th*^ and *j*^*th*^ patients,

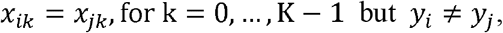

and it results in many groups with different values of risk factors. Assuming that by analyzing all patient data in a hospital for a certain period, there are *n* patients with same values of *K* risk factors,

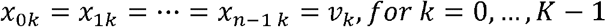

and VTE events occurred in *m* patients,

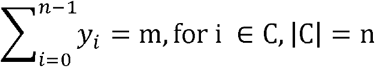

where n patients with risk factor vector v= *v*_O_,*v*_1_, …, *v*_*K*−1_) make up the group C. The *VTE risk ratio* for group C can be calculated as r= *m*/*n*.

The existence of this phenomenon limits the performance of ML model in two aspects. On one hand, most reported VTE ML models were trained using equal number of VTE and non-VTE patients, which was not consistent with distribution of the real clinical dataset. In these training set, the estimation of *VTE risk ratio* of the group C, denoted as *r*′, may be unequal to *r*, which represents the *VTE risk ratio* in the real clinical dataset. When *r*′ ≫ *r*, the ML model tends to predict patient with the risk factor vector v to be high risk and vice versa. Besides, when the training set includes more patients from the group C with risk factor vector v, the number of patients from other groups with vector *v*′ (*v*′ ≠v) tends to be less, which will influence the prediction of patients with risk factor vector *v*′. Therefore, it is unreasonable to construct the training set by including equal number of VTE and non-VTE patients. On the other hand, when the number of patients in group C is small, estimation of its real *VTE risk ratio* r may be unreliable. Especially, due to the low incidence rate of VTE, non-VTE patients are more likely to be observed in a group. If there is a group C with high *VTE risk ratio* r but only small number of non-VTE patients from it were collected, ML model based on it will predict patients of group C as low risk, which reduces model’s sensitivity.

Considering the problem of Padua model and the effect of ‘fuzzy feature’ phenomenon, to build a new VTE risk assessment model for Chinese inpatients, this study proposed a population-split-based approach as shown in **Figure 1**. This approach first split patients into different groups according to their values of risk factors, then filtered out trustless groups, and finally trained ML models in the unit of groups. It explores relationship between VTE events and combinations of risk factors by constructing different groups, rather than relationship between VTE event and risk factors, which shows advantages of robustness and good performance. Then our model was compared with Padua and multiple traditional ML models on real clinical dataset to verify its efficiency, indicating the potential to help clinicians evaluate VTE risk and guide prevention.

**Figure 1.**
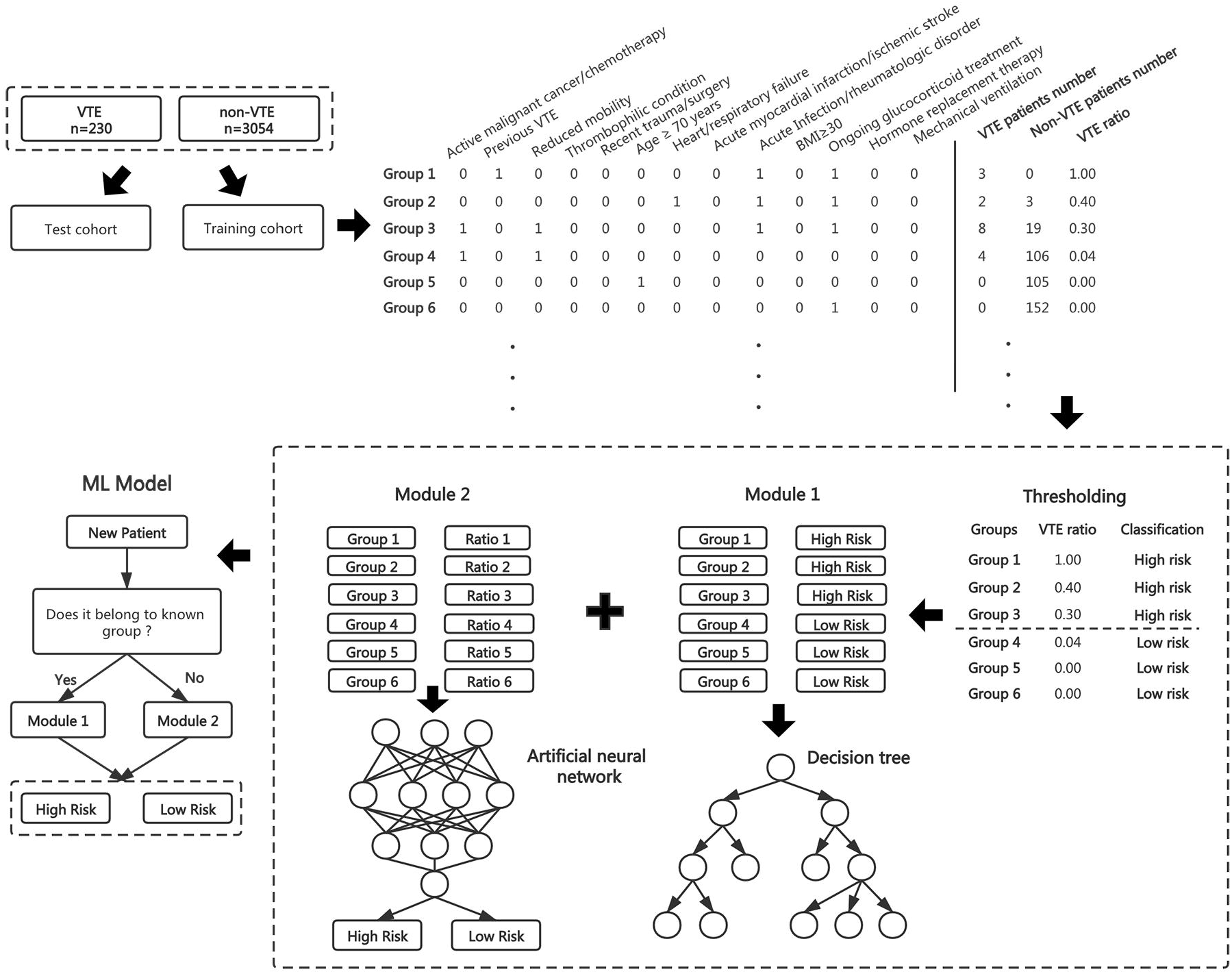
The schema of proposed VTE risk assessment ML approach. Firstly, training and test cohorts were constructed and patients in training data were split into different groups according to values of VTE risk factors. For different group *C*^(*i*)^ and *C*^(*j*)^, their corresponding feature vectors *v*^(i)^ and *v*^(*j*)^ satisfied *v*^(*i*)^ ≠ *v*^(*j*).^ Then *VTE risk ratio* was calculated in every group and groups were sorted accordingly. Next probability of distribution of patients in each group was estimated using VTE incidence rate and only groups with probability < 0.05 were saved. Based on sorted result, accumulated sensitivities and specificities were calculated for every group and groups were classified into high and low risks by thresholding, which formed a new training set based on groups. Using this training set, the proposed model consists of two modules, group-memory module for patients in known groups and group-prediction module for the unknown. Decision tree was used in group-memory module. For group-prediction module, VTE ratios for groups were used instead of high or low risk label, and artificial neural network was fitting.

## MATERIALS AND METHODS

### Population

This study analyzed inpatients who developed VTE, including DVT and PE, and partially non-VTE patients in medical department of Peking Union Medical College Hospital (PUMCH) from January 2014 to June 2016. All these inpatients from May 2016 to June 2016 were included as a test cohort for model verification and the other patients formed the training cohort. All the enrolled patients met the following inclusion/exclusion criteria: inclusion criteria: over 18 years old, hospital stay ≥72 hours; exclusion criteria: receiving anticoagulation medicine (e.g., therapeutic dose of low-molecular-weight heparin for treatment of acute myocardial infarction) other than the anticoagulation regimen for VTE diagnosed during the hospitalization.

DVT was diagnosed as the presence of intraluminal blocking or filling defects in the deep veins of the upper or lower limbs evidenced by venography or deep vein thrombogenesis illustrated by color Doppler ultrasonography. PE was diagnosed either as the presence of intraluminal blocking and/or filling defects in the pulmonary arteries by pulmonary angiography, computed tomographic pulmonary arteriography or magnetic resonance, or by radionuclide lung ventilation-perfusion scans showing multiple pulmonary segmental perfusion defects. This study was approved by the Ethics Committee of PUMCH in Chinese Academy of Medical Sciences (reference number for ethics approval: B164).

### Variable selection

The variables involved in modeling are VTE risk factors, including active cancer, previous VTE, reduced mobility, thrombophilia, recent trauma and/or surgery, age ≥70 years, heart and/or respiratory failure, acute myocardial infarction or ischemic stroke, acute infection and/or rheumatologic disorder, obesity, ongoing glucocorticoid treatment, hormone replacement therapy including estrogen or progesterone, mechanical ventilation ^11^.

### Population clustering analysis and population split

In order to learn patterns of distribution of VTE patients, population clustering analysis was performed using 16 features, including 13 VTE risk factors, Padua score, Padua high risk, and the number of non-zero risk factors. Inspired by the clustering analysis results, patients were split into different groups based on values of 13 risk factors. Denote the dataset consisted of N patients as 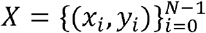, where *x*_*i*_ = (*x*_*i*1_, *x*_*i*2_, *x*_*i*3_,…, *x*_*iK*_) represented feature vector of the *i*^*th*^ patient, *x*_*ik*_ is binary variable, and *y*_*i*_ ∈ {0,1}. Now the dataset X was split into T groups 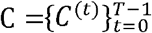, and for the *t*^*th*^ group, *n*^(*t*)^ = |*C*^(*t*)^| represented its number of patients. For every patient belonged to *C*^(*t*)^, their values of feature vector were the same 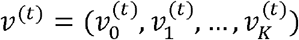. For any two groups *C*^(*t*)^ and *C*^(*q*)^, we have

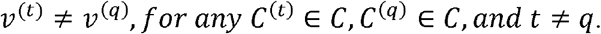

### Calculation of *VTE risk ratio* and group filtering

After splitting the dataset into T groups, *VTE risk ratio* for each group was calculated. For group *C*^(*t*)^, let 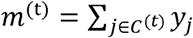 be the number of VTE patients, and its *VTE risk ratio* was 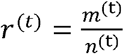. To remove groups which couldn’t represent real *VTE risk ratio*, the probability of including *m*^(t)^ VTE patients among *n*^(t)^ patients for group *C*^(t)^ was computed using the incidence rate of VTE,

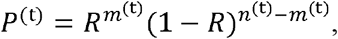

where *R* was the VTE incidence rate in whole population. Then groups with *P*^(t)^ ≥ threshold (e.g. 0.05) were filtered out and the remaining groups were saved to train model.

### Training set construction

Considering the effect of the ‘fuzzy feature’, training set was built in unit of group *C*^(*t*)^ instead of patient (*x*_*i*_, *y*_*i*_). Firstly, all saved groups were sorted by *VTE risk ratio r*^(*t*)^, number of VTE patients *m*^(*t*)^, and [-1* number of non-VTE patients *n*^(*t*)^ − *m*^(*t*)^)]. Then accumulated sensitivities and specificities were calculated from group *C*^(0)^ to *C*^(*T*−1)^. Groups after filtering were denoted as 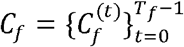 and *T*_*f*_ = |*C*_*f*_ | was the number of groups. For the *t*^*th*^ group 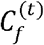, its accumulated sensitivity and specificity were

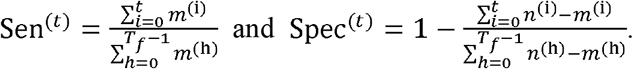

Next groups were classified into high and low VTE risks by thresholding values of Spec^(*t*)^ (e.g. 75%). The groups with Spec^(*t*)^ ≥ *threshold* were recognized as the high risk with *y*^(*t*)^ = 1, the other groups were the low risk with *y*^(*t*)^ = 0. Finally, model training set 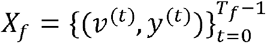 with *T*_*f*_ samples was constructed.

### Model derivation

In the training set, labels of groups were assigned based on statistical analysis, and they were regarded as the ground truth, or known knowledge. For patients from known groups 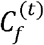, the VTE risk could be obtained simply by looking up a table consisted of all groups (*v*^(*t*)^, *y*^(*t*)^). For patients from unknown groups *C*^*unknown*^,

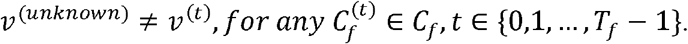

Reasonable and accurate VTE risk prediction for these unknown patients was needed based on results of known groups, which was the goal of training a ML model.

Thus, the proposed VTE risk assessment model consisted of two modules, the group-memory module for patients from known groups and group-prediction module for patients from unknown groups. For group-memory module, a decision tree model was used to record all pairs (*v*^(*t*)^, *y*^(*t*)^) from *X*_*f*_, and contributions of risk factors could be analyzed by comparing feature weights. For group-prediction module, an artificial neural network (ANN) was used to fit the relationship between feature vector of group *v*^(*t*)^ and *VTE risk ratio r*^(*t*)^. By comparing goodness of fit, the optimal ANN with the highest ^2^ was selected, and patient with predicted VTE ratio ≥ 0.5 was recognized as the high VTE risk.

### Model evaluation and comparison

To verify the proposed model’s efficiency, five traditional ML models including SVM,RF, GBDT, LR ^18^, and XGBoost ^19^, and Padua model, were compared. Five ML models were trained in the popular method, on the same training patients as the proposed model, and considering that the number of non-VTE patients were larger than the number of VTE, non-VTE patients equal to the number of VTE patients were randomly selected to construct 1:1 training set. For five ML models, 10-fold cross validation was used and the optimal ML models were chosen with the highest Youden index ^20^. Model’s sensitivity, specificity, and Youden index were computed to evaluate their predictive validity, and the training process was repeated five times to calculate mean values and standard deviations.

## RESULTS

### Characteristics of distribution of VTE patients

230 VTE patients and 3054 non-VTE patients were included in this study. Clustering analysis with 13 VTE risk factors and 3 Padua-score-related features on these patients, (**Figure 2**) showed that VTE patients didn’t get together and were scattered among non-VTE patients. The distribution of VTE patients with a Padua score ≥ 6 points (accounting for 49.13% of overall VTE patients) is more intensely concentrated, while VTE patients with Padua score <6 points were poorly characterized by the Padua model and distributed over a wide area (accounting for 50.87% of overall VTE patients). 14.35% VTE patients had a Padua score under 4 points and were stratified incorrectly as low risk. In addition, 85.87% of the high-risk patients recognized by the Padua were non-VTE patients.

**Figure 2.**
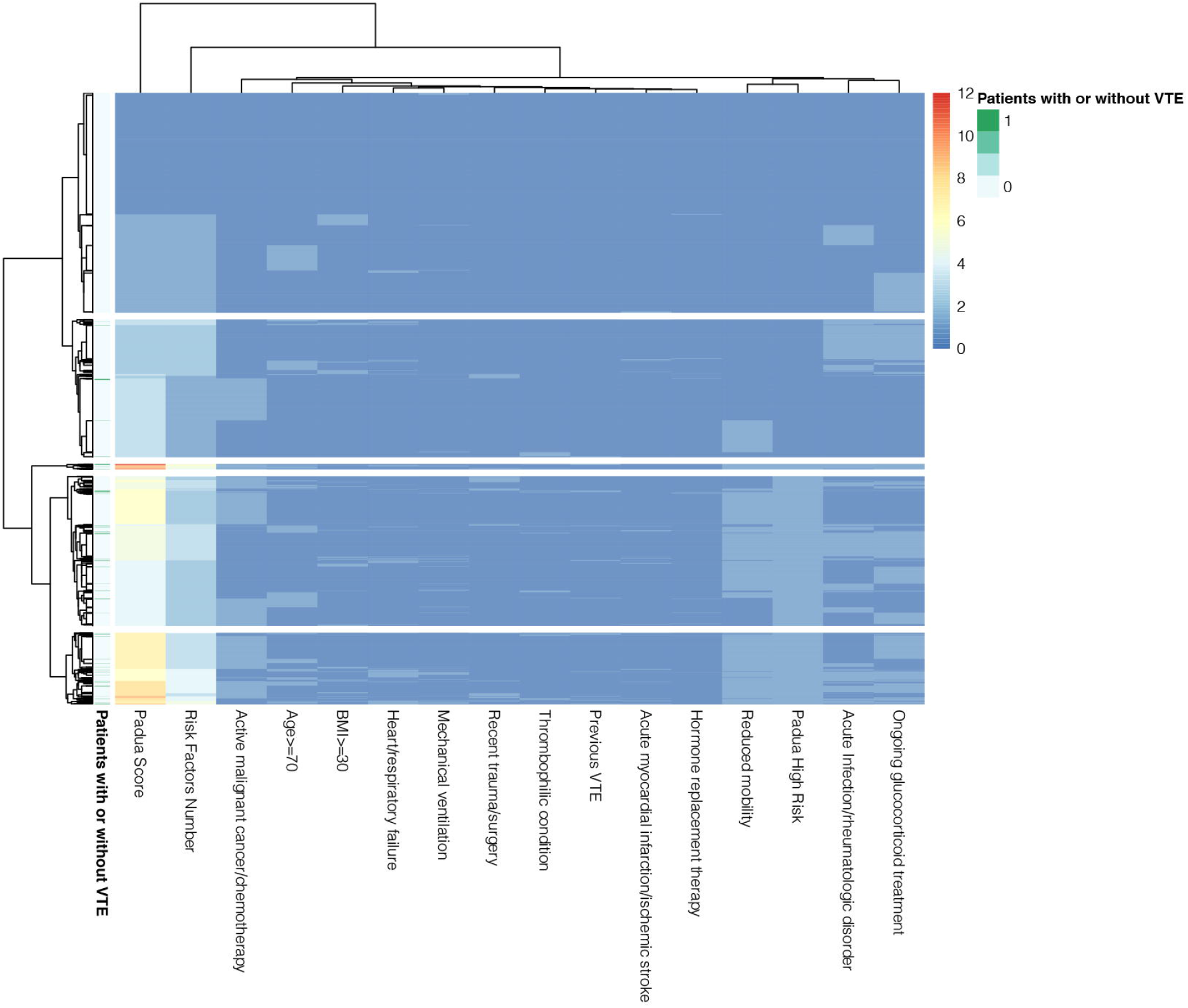
Population clustering analysis on inpatients from PUMCH. The clustering analysis with 13 VTE risk factors and 3 Padua-score-related features (Padua score, the number of VTE risk factors, and Padua high risk) was conducted on 3284 inpatients from PUMCH including 230 VTE and 3054 non-VTE patients. Each row represented a patient with (labeled with dark green color) or without (labeled with light green color) VTE in this heatmap. Features listed in the columns were labeled dark blue color as a lower value and red color as a higher value.

Inspired by the clustering analysis results, patients with same values of risk factors were grouped and the ratio of VTE patients were calculated. Some representative groups were shown at **Table 2**. It could be seen that there were VTE and non-VTE patients with identical values of risk factors and different groups had distinct *VTE risk ratios*, which proved the existence of ‘fuzzy feature’. Four groups in **Table 2** had both VTE and non-VTE patients and there were two groups with only non-VTE patients. The 4^*th*^ group with feature vector v = (0,0,0,0,…, 0) had more patients than any other groups, which meant that most of patients didn’t have non-zero VTE risk factor. The 3^*th*^ group with v = (0,0,1,0,…,0) was the second largest group, which showed that there were many patients with just one risk factor, the reduced mobility. The 1^*th*^ group with v = 0,0,1,0,…, 0) had more VTE than the non-VTE patients, but the 2^*th*^ group with v = (1,0,1,0,…,0) had less VTE than the non-VTE patients.

**Table 2.** Representative groups and their VTE ratio in PUMCH data.

| Group Index | Feature vector of group | VTE Patients | Non-VTE Patients | VTE Ratio |
| --- | --- | --- | --- | --- |
| 0 | 0,1,0,0,0,0,0,0,1,0,1,0,0 | 3 | 0 | 100% |
| 1 | 0,0,1,0,0,1,1,0,1,0,0,0,0 | 4 | 2 | 66.67% |
| 2 | 1,0,1,0,0,0,0,0,1,0,1,0,0 | 8 | 30 | 21.05% |
| 3 | 0,0,1,0,0,0,0,0,0,0,0,0,0 | 3 | 168 | 1.75% |
| 4 | 0,0,0,0,0,0,0,0,0,0,0,0,0 | 2 | 637 | 0.31% |
| 5 | 1,0,1,0,0,0,0,0,1,1,0,0,0 | 0 | 2 | 0% |
| 6 | 1,0,1,0,1,0,0,0,0,0,0,0,0 | 0 | 13 | 0% |

### Mean predictive validity of VTE risk assessment models

The training data consisted of 189 VTE patients and 1531 non-VTE patients, while test data included 41 VTE patients and 1523 non-VTE patients. Mean values of sensitivities and specificities of five ML models, Padua and the proposed model on all training and test patients were listed in **Table 3**. Compared to the result of Padua model, generally five ML models had relatively higher specificities but lower sensitivities. Within five ML models, RF was the only model with sensitivities > 0.80 on both training and test data and average performance of specificities (>0.80) of XGBoost were the best. There was no model with both higher sensitivity and specificity than the Padua among five ML models. However, on the training data, the proposed model achieved advantages on both sensitivity and specificity over the Padua. On the test data, mean values of sensitivities of the proposed model were very similar with the Padua and specificities of the proposed were higher. In addition, standard deviations of predictive validity of the proposed model were far less than the five ML models.

**Table 3.** Comparison of mean predictive validity of five ML models, Padua and proposed model.

| Model Name | Training set |  | Test set |  |
| --- | --- | --- | --- | --- |
|  | Sensitivity | Specificity | Sensitivity | Specificity |
| SVM | 0.7894±0.0220 | 0.7240±0.0431 | 0.8341±0.0420 | 0.7032±0.0424 |
| RF | 0.8413±0.0150 | 0.7757±0.0138 | 0.8195±0.0452 | 0.7349±0.0156 |
| GBDT | 0.7883±0.0404 | 0.8135±0.0394 | 0.8146±0.0396 | 0.7825±0.0441 |
| LR | 0.7397±0.0304 | 0.7960±0.0230 | 0.8195±0.0293 | 0.7869±0.0230 |
| XGBoost | 0.7524±0.0316 | 0.8328±0.0185 | 0.8293±0.0218 | 0.8064±0.0239 |
| **Padua** | **0.8466** | **0.6127** | **0.9024** | **0.6330** |
| **Our** | **0.8995±**<br>**1.110E-16** | **0.6741±0.0056** | **0.9024±**<br>**1.110E-16** | **0.6453±0.0033** |

Values of sensitivity and specificity were represented with ‘mean value ± standard deviation’. The model training process was repeated five times to calculate the predictive validity. Note that sensitivities and specificities on training process were computed using all patients from training data.

### The optimal predictive validity of VTE risk assessment models

Further, the optimal ML models were selected by cross validation for five ML models and the proposed model, and their performances on training and test data were shown in **Table 4**. In general, patterns of ML models were consistent with the results in **Table 3**. Five ML models had higher specificities by sacrificing the sensitivities. Value of Youden index of the optimal proposed model was not as high as the RF, GBDT and XGBoost, but better than the SVM, LR, and Padua. However, the optimal proposed model had the highest sensitivity and its specificity was better than the Padua on both training and test data, which verified its excellent consistency.

**Table 4.** Comparison of predictive validity of the optimal ML models, proposed model and Padua.

| Model<br>Name | Training set |  |  | Test set |  |
| --- | --- | --- | --- | --- | --- |
|  | Sensitivity | Specificity | Youden | Sensitivity | Specificity |
| SVM | 0.8042 | 0.7511 | 0.5554 | 0.8292 | 0.7104 |
| RF | 0.8307 | 0.7975 | **0.6282** | 0.8780 | 0.7643 |
| GBDT | 0.8148 | 0.8223 | **0.6372** | 0.8780 | 0.7787 |
| LR | 0.7725 | 0.7864 | 0.5589 | 0.8537 | 0.7708 |
| XGBoost | 0.7883 | 0.8302 | **0.6185** | 0.8537 | 0.7919 |
| **Padua** | **0.8466** | **0.6127** | **0.4593** | **0.9024** | **0.6330** |
| **Our** | **0.8995** | **0.6786** | **0.5781** | **0.9024** | **0.6494** |

The optimal ML models and proposed model were selected by maximizing the value of Youden index on training data. Note that metrics of predictive validity on training process were computed using all patients from training data.

## DISCUSSION

In this study, we proposed a new VTE risk assessment model which split the population into groups based on values of risk factors, and established group-memory and group-prediction modules respectively in order to consider the effect of ‘fuzzy feature’ and describe VTE patients’ characteristics better. By comparing with five traditional ML models and Padua model on patients from PUMCH, effectiveness of our proposed model was validated and it showed better robustness than traditional ML models trained on equal number of VTE and non-VTE patients. The proposed model was the only one which showed advantages on both sensitivity and specificity over Padua model.

For five ML models trained on equal number of VTE and non-VTE patients, results in **Table 4** were calculated by using default threshold 0.5 for predictive probability to classify patients into the high or low risks. Due to the fact that sensitivity and specificity of model can be different by changing values of thresholds, the relationship between predictive validity, namely sensitivity and specificity, and the threshold was explored further and plotted at **Figure 3**. RF, GBDT, and XGBoost were selected typically because they achieved higher values of Youden index than the proposed model. From **Figure 3** it could be seen that, on training and test data, for GBDT and XGBoost, there was not a threshold that had higher sensitivity and specificity simultaneous than the proposed model. For RF, thresholds with better predictive validity than the proposed model only existed on test data. In summary three ML models couldn’t obtain higher sensitivities and specificities than the proposed by changing predictive thresholds, which proved our model’s efficiency again.

**Figure 3.**
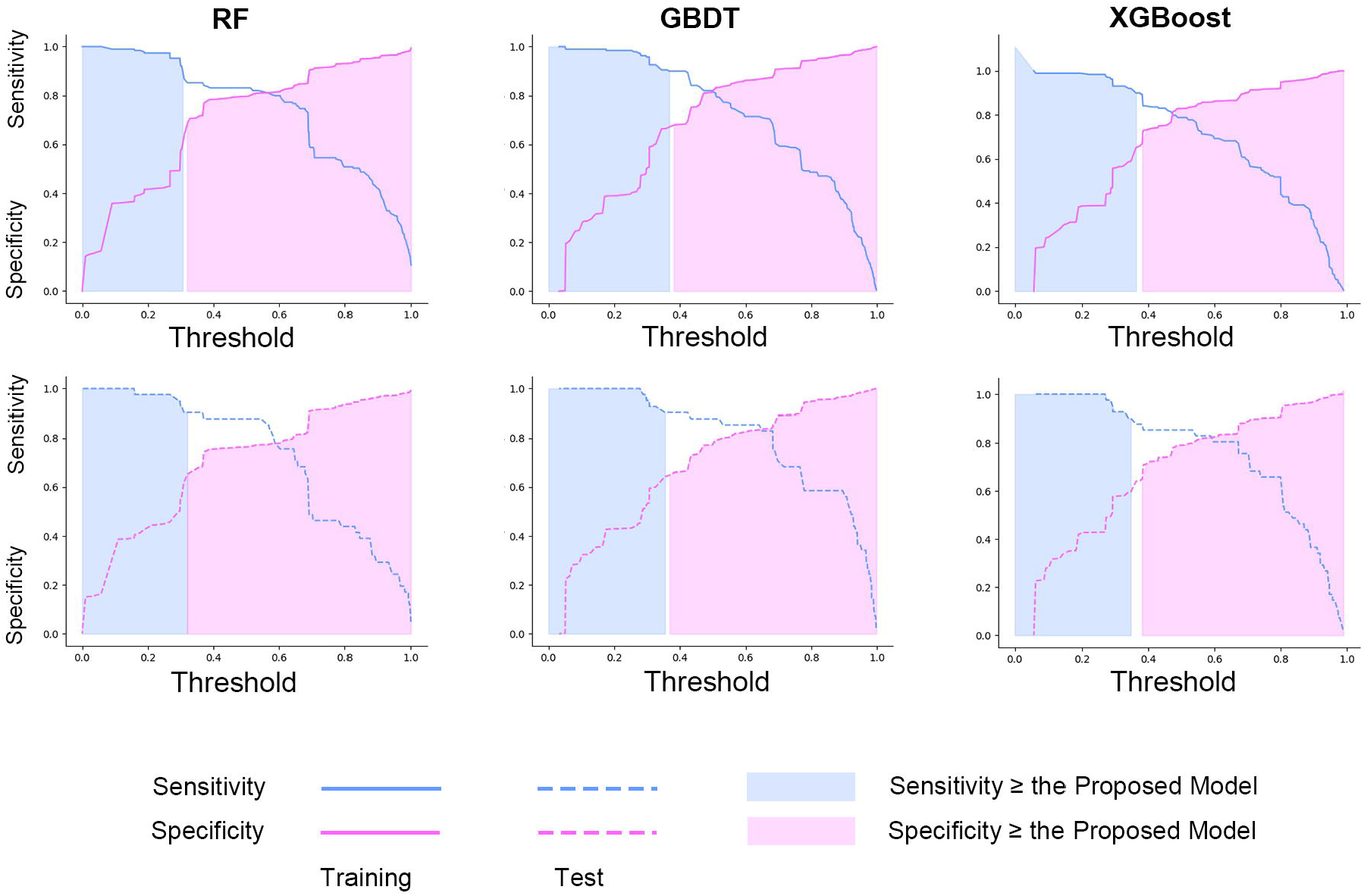
Changing of sensitivities and specificities of three ML models with the increasing of predictive thresholds. 3 ML models with higher Youden index (RF, GBDT and XGBoost) than the proposed model were selected. Results in training and test data were shown in upper and lower figures respectively. For each figure, thresholds with higher specificities than the proposed model were marked with pink, and with higher sensitivities were marked with blue.

One notable result from **Table 3** was that standard deviations of sensitivities and specificities of five ML models trained on equal number of VTE and non-VTE patients were larger than our proposed model, which showed that ML models, lacked robustness. Due to the neglect of ‘fuzzy feature’, within training set, randomly selected non-VTE patients would disturb the estimation of *VTE risk ratio*s of groups, which lead to instability of model’s performance. To elaborate the influence of ‘fuzzy feature’ on ML models’ predictive performance, changing of sensitivities and specificities of ML models by strengthening the ‘fuzzy feature’ was visualized in **Figure 4**. For group *C*^(*t*)^ ∈ *C*_*fuzzy*_ which included less VTE patients than non-VTE patients, namely *VTE risk ratio* 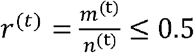, by increasing the number of non-VTE patients of group *C*^(*t*)^ in the training set, VTE ratio *r*^(*t*)′^ could be changed from >0.5 to ≤ 0.5. When *r*^(*t*)′^ ≤ 0.5, the ML model tended to predict patients from group *C*^(*t*)^ as the low VTE risk, which would affect model’s performance. It could be seen that with the increasing of ratio of groups with *r*^(*t*)′^ ≤ 0.5 belonged to *C*^(*t*)^ in training set, sensitivities and specificities of three ML models varied dramatically. Therefore, taking the ‘fuzzy feature’ into account was crucial to model’s robustness. Actually, “fuzzy feature” is very common in medical area, since the incidence of most diseases is limited. This method can be widely used in many aspects, especially disease screening or risk prediction.

**Figure 4.**
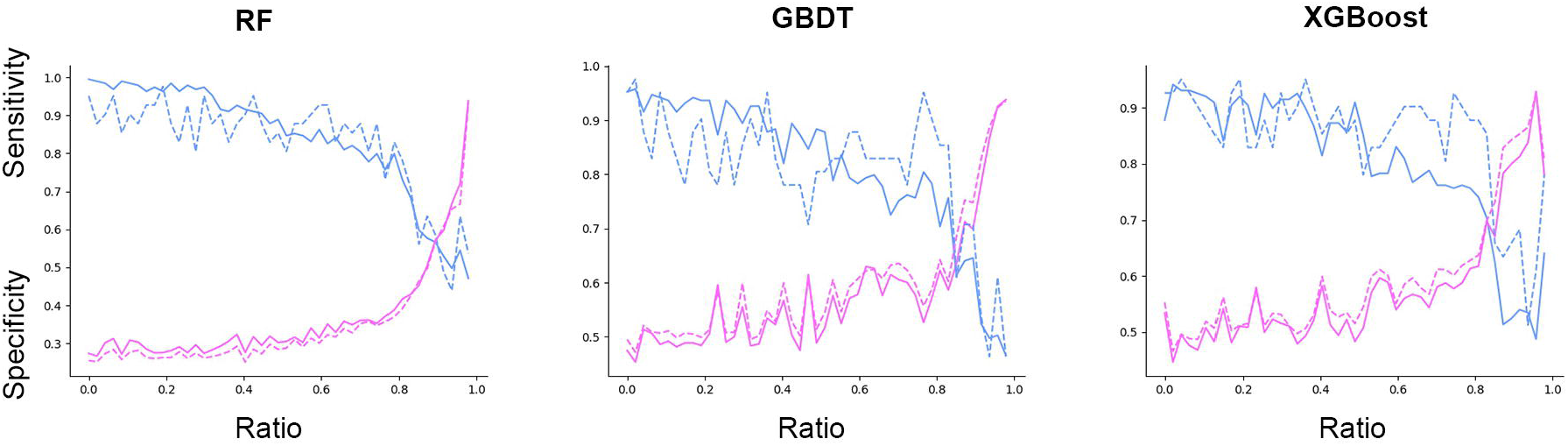
Variation of sensitivities and specificities of three ML models by strengthening the effect of ‘fuzzy feature’. RF, GBDT and XGBoost were selected to show the influence of ‘fuzzy feature’. For all patients in training data, groups set *C*_*fuzzy*_ = {*C*^(*t*)^}where *VTE risk ratio* 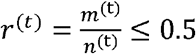 was recognized and groups in *C*_*fuzzy*_ were sorted according to patients’ sizes *n*^(t)^. During the process of constructing 1:1 (VTE: non-VTE) training set, the varying of number of included non-VTE patients *n*^(t)′^ − *m*^(t)′^ from *C*^(*t*)^ could change VTE ratio *r*^(*t*)′^ from > 0.5, *when* 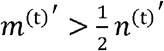, to ≤ 0.5,when 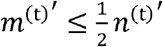, which would influence the prediction of ML models on patients in *C*^(*t*)^. By proportionally setting VTE ratios of groups in *C*_*fuzzy*_ into *r*^(*t*)′^ ≤ 0.5 manually, namely including more non-VTE patients than VTE patients from *C*^(*t*)^ (*n*^(t)′^ − *m*^(t)′^ ≥ *r*^(t)′^) in the training set, influence of ‘fuzzy feature’ on models’ predictive validity was visualized. *m*^(t)^, the number of VTE patients in group *C*^(*t*)^; *n*^(*t*)^, the number of all patients in *C*^(*t*)^.

Currently our study still needs to be improved in several aspects. Firstly, due to the low incidence of VTE, the sample size of this research center is still limited. To assess statistical differences of predictive validities between the proposed model and Padua model, studies with larger sample sizes are still required. Multi-center and prospective researches are also needed to validate and promote the model further. Secondly, with increasing number of VTE samples, the deep learning methods ^21,22^ maybe can replace the ANN model to further improve our model.

## CONCLUSION

Based on population clustering analysis and the Padua model, by considering the effect of ‘fuzzy feature’, this study proposes a new VTE risk assessment model using data from Chinese medical inpatients. The model shows a promising performance in VTE risk prediction, which may help clinicians stratify VTE risk and guide prevention efficiently.

## Data Availability

All data produced in the present study are available upon reasonable request to the corresponding author

## Author Contributions

**Xin Wang & Yu-Qing Yang:** Conceptualization, Methodology, Formal analysis, Writing - original draft. **Xin-Yu Hong & Si-Hua Liu:** Investigation, Data curation. **Jian-Chu Li & Ting Chen:** Supervision, Writing - review & editing. **Ju-Hong Shi:** Resources, Supervision, Project Administration, Funding acquisition, Writing - review & editing.

## Acknowledgement

This work is supported by the Chinese Academy of Medical Sciences Fundamental Research Funds (2019XK320044).

## Summary Points

What was already known on the topic:

- For the VTE risk prediction of Chinese inpatients, the performance of Padua model, which is recommended for inpatients by the American College of Chest Physicians, is inferior.
- Multiple ML models were proposed to predict VTE risk but they lack robustness and cannot achieve higher sensitivity and specificity over the Padua simultaneously.

What this study adds to our knowledge:

- An important statistical phenomenon, ‘fuzzy feature’, which is very common in medical area for diseases with low incidence, is elaborated.
- Neglecting the influence of ‘fuzzy feature’, previous ML models achieved unstable VTE risk prediction results.
- A new VTE risk assessment ML model is proposed by considering the effect of ‘fuzzy feature’. Its predictive performance is better than the Padua and its robustness is higher than traditional ML models.

